# Vaccination and testing as a means of ending the COVID-19 pandemic: comparative and statistical analysis

**DOI:** 10.1101/2022.06.16.22276531

**Authors:** Igor Nesteruk

## Abstract

**Background:** Record numbers of new cases and deaths registered in Japan and European countries in early 2022 once again proved that existing vaccines cannot stop the new infections and deaths caused by SARS-CoV-2 and aroused new questions about methods of overcoming the pandemic.

**Aim of the study:** to compare the pandemic waves in Japan, Ukraine, USA, Hong Kong, mainland China, European and African countries in 2020, 2021, 2022 and to investigate the influence of testing and vaccination levels.

**Methods:** The smoothed daily numbers of new cases and deaths per capita, the ratio of these characteristics and the non-linear correlation with the tests per case ratio were used.

**Results:** As in other countries, the deaths per case ratio in Japan decreases with the increase of the vaccination level. Non-linear correlation revealed, that the daily number of new cases drastically decreases with the increase of the tests per case ratio.

**Conclusions:** Increasing the level of testing (especially for people who may have contact with infected persons) and adhering to quarantine restrictions for the entire population, including vaccinated people, may be recommended to end the pandemic.

## Introduction

The first studies of the effectiveness of SARS-CoV-2 testing appeared in 2020 immediately after the pandemic outbreak (see, e.g., [1-15]). The effectiveness of testing and isolation in reducing the basic reproduction number was investigated in recent paper [16] based on mathematical modeling. In 2021, with the advent of a sufficient number of vaccinated people, it became possible to study the impact of vaccination levels on the COVID-19 pandemic dynamics (see, e.g. [17-25]). Constant changes in its dynamics caused by changes in quarantine conditions and algorithms for testing and vaccination, the emergence of new strains require updating and rethinking the results of previous studies. To compare the pandemic situation in different countries with different sizes of their population the relative characteristics can be used: e.g., the daily numbers of new COVID-19 cases per capita (DCC), deaths per capita (DDC), and tests per capita (TC)

Usually the values of DCC, DDC and TC are very random and show some weekly periodicity. For example, the World Health Organization reports only weekly numbers of new cases and deaths after summer 2020, [26]. COVID-19 Data Repository by the Center for Systems Science and Engineering (CSSE) at Johns Hopkins University (JHU) provides the daily figures of DCC, DDC, TC and calculates also their smoothed values by averaging numbers corresponding to a fixed day and six previous days [27]. We will take these values for our analysis without using the smoothing procedure proposed in [28-30].

Since we are already in the third year of the pandemic, it is reasonable to compare the DCC, DDC and DDC/DCC numbers in 2020, 2021, and 2022 in order to find some seasonal trends. Since the influence of vaccinations in 2020 and early 2021 can be neglected, the comparison of the COVID-19 pandemic dynamics for the later period enables us to reveal some influences of vaccination levels.

In [31] the DCC values for Argentina, Brazil, India, South Africa, Ukraine, EU, the UK, USA and the whole world for the period from March to August 2020 were compared with the corresponding values in 2021. The DCC, DDC, DDC/DCC values and vaccination levels in the same countries and Australia have been compared for the period from September to January in 2020-2021 and 2021-2022, [22]. In this paper we will focus on the DCC, DDC, DDC/DCC values registered in Japan in 2020, 2021, 2022 and will try to find their links with the percentage of fully vaccinated people (VC) and busters (BC), the daily numbers of tests per capita TC and the daily tests per case ratio (DTC).

The pandemic dynamics in European and African countries; Japan, USA and some other countries will be compared. The non-linear regression will be applied to find links between the DCC and DTC values.

## Materials and Method

We will use the smoothed daily numbers of new cases (DCC, per million), deaths (DDC, per million) and tests (TC, per thousand) registered by JHU, [27] in 2020, 2021 and 2022. We will also use the accumulated percentages of fully vaccinated persons (VC, in 2021 and 2022) and persons who received booster doses of vaccines (BC, in 2022) listed in [27]. Since JHU often updates its data sets, we will specify the versions of its file used in different cases.

To find links between DCC values and the daily tests per case ratio DTC=1000*TC/DCC we will use the non-linear correlation,

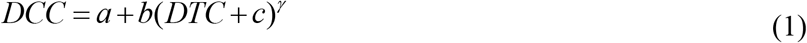

At *γ* =1 relationship (1) reduces to a linear one and can be rewritten as follows:

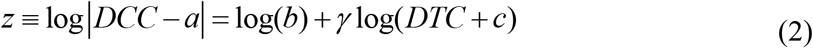

Then for new random variables *z* and *w* ≡ log(*DTC* + *c*) we will have a linear regression. The constant parameters *γ*, log(*b*) and corresponding best fitting lines can be found with the use of standard formulas for the linear regression [32] for every fixed values of constant parameters *a* and *c*.

We will use also the F-test for the null hypothesis that says that the proposed linear relationship (2) fits the data sets. The experimental values of the Fisher function can be calculated with the use of the formula:

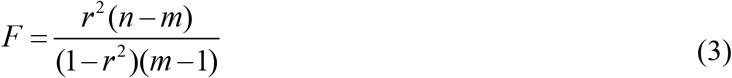

where *n* is the number of observations (number of countries and regions taken for statistical analysis); *m*=2 is the number of parameters in the regression equation, [32]. The corresponding experimental values *F* have to be compared with the critical values *F*_*C*_ (*k*_1_, *k*_2_) of the Fisher function at a desired significance or confidence level (*k*_1_ = *m* −1, *k*_2_ = *n* − *m*, see, e.g., [33]). If *F*/*F*_*C*_ (*k*_1_, *k*_2_) < 1, the null hypothesis is not supported by the results of observations. The highest values of *F*/*F*_*C*_ (*k*_1_, *k*_2_) correspond to the most reliable hypotheses (see, e.g., [34]). In the case of non-linear regression (2) the values of additional parameters *a* and *c* must be chosen in order to ensure the maximum of the correlation coefficient *r* or the highest values of to the parameter *F*/*F*_*C*_ (*k*_1_, *k*_2_) (similar to the parameter identification methods used in [29]).

## Results

Figs. 1 and 2 illustrate the differences in epidemic characteristics in Japan in 2020 (dashed lines), 2021 (solid lines) and 2022 (bold solid lines). We have used the figures corresponding to the version of JHU file published on May 26, 2022, [27]. Red lines in Fig. 1 represent the soothed daily numbers of cases and display 6 distinct epidemic waves corresponding to the maximum figures of DCC. The maximum values of new daily cases were registered in May-June (2020, 2021), August (2020, 2021) and January-February (2021, 2022). Thus, we can speak about some seasonal influences on the pandemic dynamics. The highest DCC values were registered in February 2022 despite very high levels of vaccination (see red markers in Figs. 1 and 2).

**Fig. 1.**
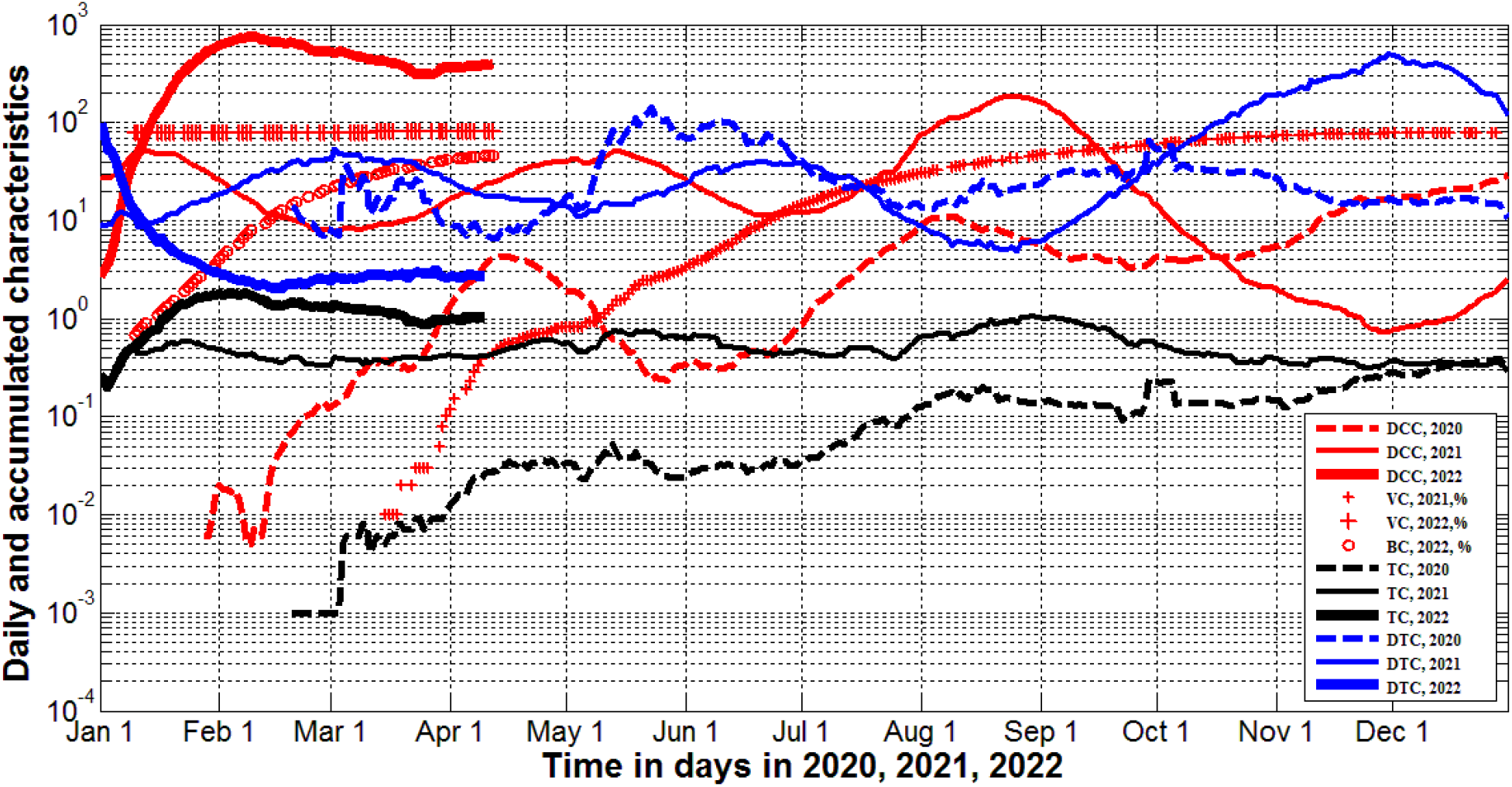
Pandemic dynamics in Japan in 2020, 2021, and 2022. Averaged daily number of new cases (DCC, red lines) and tests (TC, black) per capita, vaccination levels (VC and BC, red markers) and daily tests per case ratio (DTC=1000*TC/DCC, blue). Dashed lines correspond to 2020, solid lines – to 2021 and solid bold lines - to 2022. Small red “crosses” represent the percentage of fully vaccinated persons in 2021, large ones – in 2022; “circles” show the percentage of boosters.

**Fig. 2.**
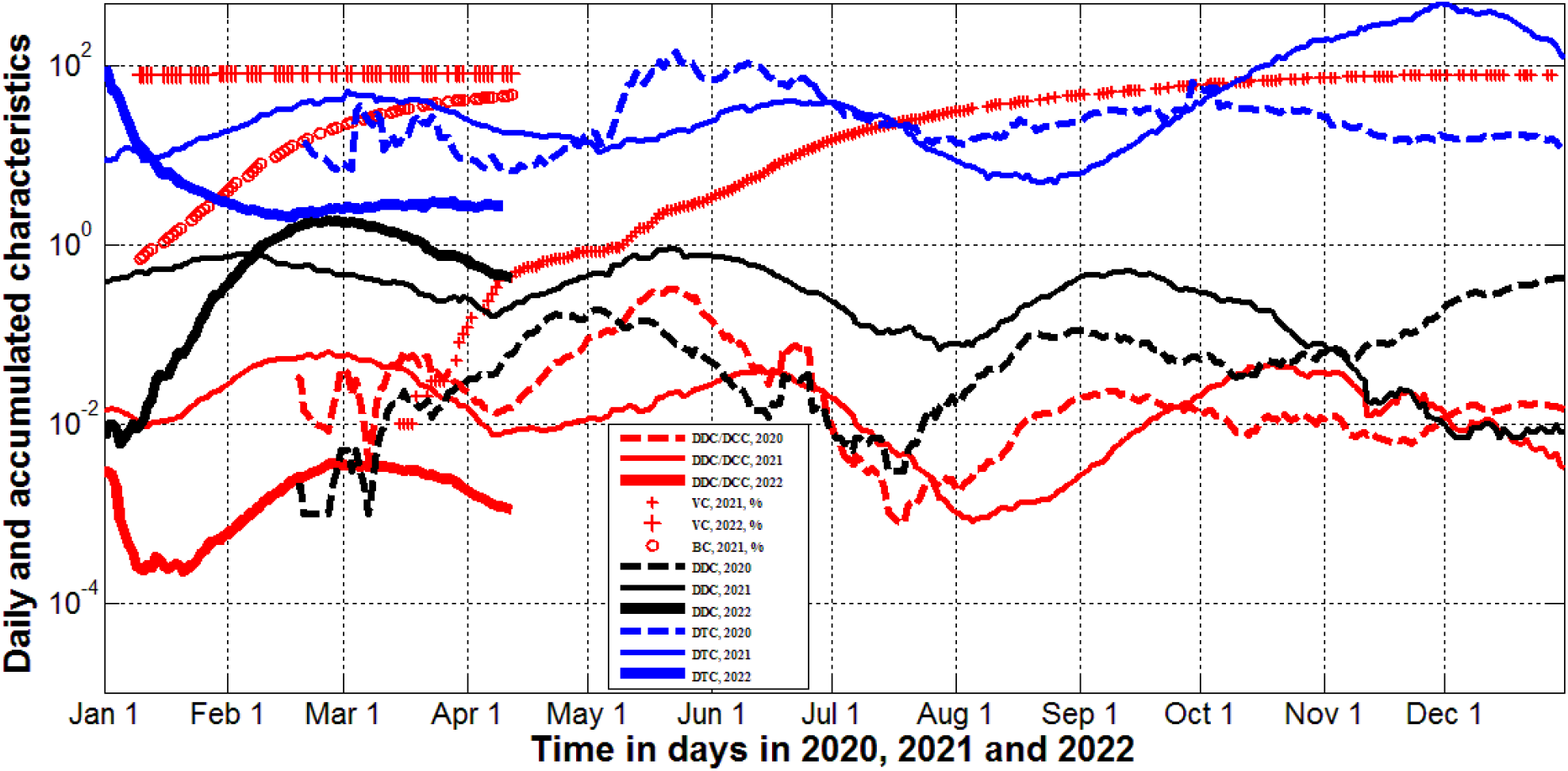
Pandemic dynamics in Japan in 2020, 2021, and 2022. Averaged daily number of new deaths (DDC, black lines), mortality rate (DDC/DCC, red lines), vaccination levels (VC and BC, red markers) and daily tests per case ratio (DTC=1000*TC/DCC, blue lines). Dashed lines correspond to 2020, solid lines – to 2021 and solid bold lines - to 2022. Small red “crosses” represent the percentage of fully vaccinated persons in 2021, large ones – in 2022; “circles” show the percentage of boosters.

Similar pandemic waves caused by the Omicron strain were registered in January 2022 in the highly vaccinated UK, USA and EU (see, e.g., [23]). Moreover simple statistical analysis (based on the JHU datasets corresponding to February 1, 2022) revealed the increase in DCC values with the growth of the percentage of fully vaccinated people in European countries and worldwide [24]. The same analysis based JHU datasets reported on September 1, 2021 revealed no statistically significant correlation between DCC and VC values, [25]. These changes in the influence of vaccination level can be explained by lifting the quarantine restrictions for vaccinated people in European countries and USA.

Japan does not reduce quarantine restrictions for vaccinated people. The solid red line, small red “crosses’ in Fig. 1 and blue “circles” in Fig. 3 demonstrate that daily number of new cases diminished with the increase of the vaccination level in September-December 2021. But the trend has changed in 2022 (compare blue “crosses” and “circles” in Fig. 3). Sharp increase in DCC values despite very high vaccination level in Japan demonstrated ones more that existing vaccine cannot stop the appearance of new cases.

**Fig. 3.**
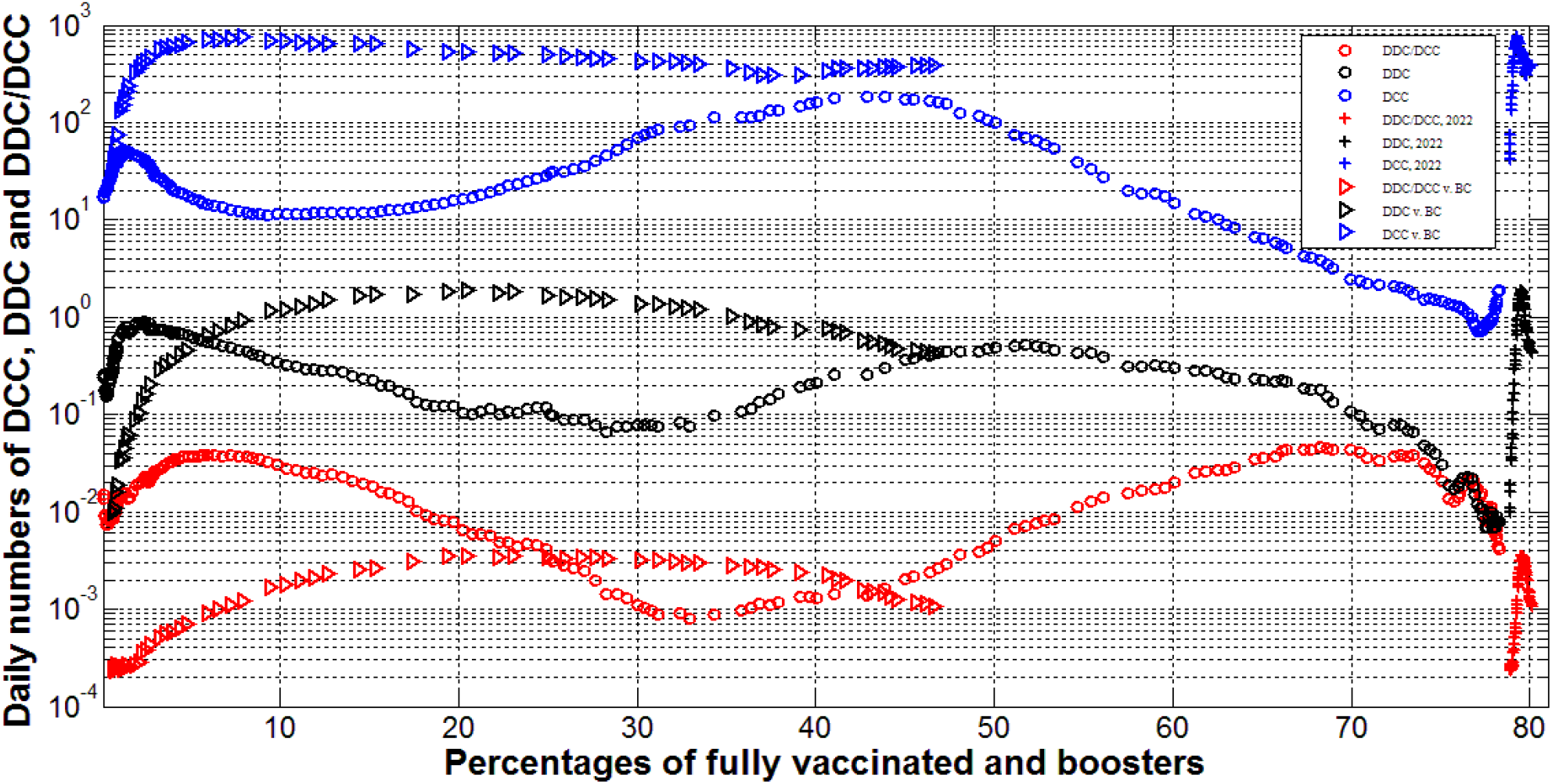
Pandemic dynamics in Japan in 2021 and 2022. Averaged daily number of new cases (DCC, blue), deaths (DDC, black) and mortality rate (DDC/DCC, lines) versus percentage of fully vaccinated persons (“circles” for 2021, “crosses” for 2022) and versus percentage of boosters (“triangles” for 2022).

Nevertheless, vaccination can reduce mortality. The decrease of DDC and DDC/DCC values with the growth of VC was demonstrated for European countries by linear regression analysis performed in [24]. In the case of Japan we see the record increase of daily numbers of deaths DDC in 2022 with the maximum level corresponding to the end of February 2022 (approximately two weeks later when the maximum of DCC was achieved, see the black bold line in Fig. 2 and the red bold line in Fig. 1). Thus, very high VC figures have not reduced daily numbers of deaths in Japan. Comparison of black “circles” and “crosses” in Fig. 3 also supports this conclusion.

The dynamics of the mortality rate DDC/DCC in Japan is more similar to the case of European countries. In January-March 2022 it was much lower than in the same period in 2021 (compare the solid bold red and the solid red lines in Fig. 2 or red “crosses” and “circles” in Fig. 3). This fact can be related to higher vaccination level in 2022. Nevertheless, very low DDC/DCC values were also registered in August 2021 (see the solid red line in Fig. 2 and red “circles” in Fig. 3) despite the low level of VC. Similar low mortality rates were registered also in July 2020 (see the dashed red line in Fig. 2) before starting the vaccinations. We can explain this fact by the influence of the seasonal factors. The probability of deaths caused by SARS-CoV-2 seems to be lower in the summer.

The increasing percentage of boosters BC have not reduced the DCC, DDC and DDC/DCC values in 2022 (see blue, black and red “triangles” in Fig. 3). Probably, we will see some positive trends later. As of February 1, 2022, the DCC values increased, but DDC and DDC/DCC figures decreased with the growth of BC for European countries, [24].

Since the existing vaccines can reduce only the probability of death for an infected person, of particular relevance is the question of methods that could stop or minimize the number of new cases. Daily numbers of new cases per million versus tests per case ratio are shown in [15] for different countries as of January 17, 2021. Despite the rather chaotic nature of the data, we can see a clear trend of decreasing the number of new cases with an increase in the tests per case ratio. Linear regression analysis applied for the datasets corresponding to European and some other countries (including Japan) as of February 1, 2022 showed that the increase of the tests per case ratio TC=1000*DTC/DCC reduces the daily numbers of new cases, [24].

Black lines in Fig. 1 represent the averaged daily numbers of tests per capita DTC in Japan. These values were more or less stable in 2021 (see the solid black line). In 2020 the level of testing was much lower (the dashed black line), in 2022 it became slightly higher (the solid bold black line). No significant correlations with the daily numbers of new cases (blue lines) are visible.

In comparison, the maximal values of the tests per case ratio (see blue lines in Figs. 1 and 2) correspond to the minimal figures of DCC (see red lines in Fig. 1) and DDC (black lines in Fig. 2). Similar correlation was revealed in [15], where the test positivity rate (a characteristics inverse to the tests per case ratio) was used. To make these correlations more visible, values of DCC, DDC and DDC/DCC are plotted versus the daily tests per case ratio DTC=1000*TC/DCC in Fig. 4. Blue and black markers illustrate the decreasing of DCC and DDC values with the growth of the tests per case ratio. The DDC/DCC values are more scattered and show no decreasing trend (see red markers).

**Fig. 4.**
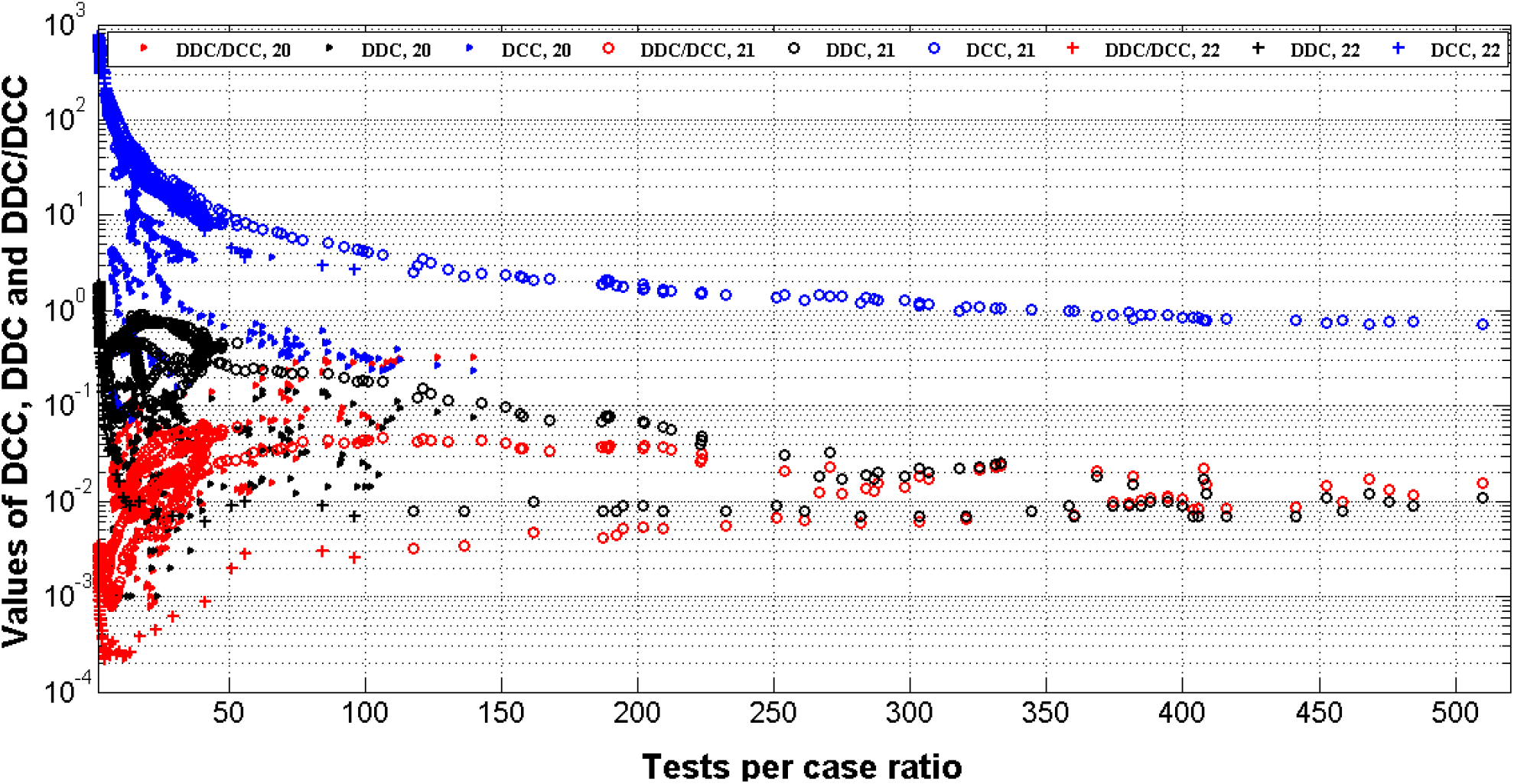
Pandemic dynamics in Japan in 2020, 2021, and 2022. Averaged daily number of new cases (DCC, blue), deaths (DDC, black) and mortality rate (DDC/DCC, red) versus daily tests per case ratio (DTC=1000*TC/DCC) in 2020, 2021 and 2022. “Triangles” correspond to 2020, “circles” - to 2021, “crosses” - to 2022.

We have applied the nonlinear correlation (1) to find the links between DCC values and the daily tests per case ratio. The statistical analysis was performed separately for Japan and USA and for three years: 2020, 2021 and 2022. The results of calculations are shown in Table 1 and Figs. 5, 6. The optimal value of parameter *c* = - 1 for all cases except the United States in 2022, for which *c* = -1.09. It can be seen that there are statistically significant correlations for both countries, since *F*/*F*_*C*_ (*k*_1_, *k*_2_) >1 for all the cases shown in Table 1. Even in 2020 (when the data was rather scattered in Japan, Ukraine, USA, and Hong Kong, see Figs. 5 and 6) non-linear correlation (1) was supported by the results of observations at high confidence level 0.001. In 2021 and 2022 the experimental points are very close to the best fifing lines (solid for Japan and dashed for USA). The experimental datasets for Ukraine in 2021 and 2022 are very close to the best fitting lines for Japan in 2021 and 2022 (compare blue and red solid lines with blue and red “triangles” in Fig. 6). In 2020, the number of cases in Ukraine were higher than in Japan at fixed values of the tests per case ratio (compare the black solid line and black triangles in Fig. 6). The number of cases in USA in 2021 and 2022 were approximately 6-8 times higher than in Japan and Ukraine at the same values of the daily tests per case ratio (see Fig. 6). This fact probably reflect the lower level of social distancing in USA.

**Table 1.** Optimal values of parameters in eq. (1), correlation coefficients and the results of Fisher test applications for Japan and USA in 2020, 2021 and 2022.

| Year, Country | Number of observations<br>$n$ | Correlation coefficient<br>$r$ | Optimal values of parameter<br>$a$<br>in eq. (1) | Optimal values of parameter<br>$b$<br>in eq. (1) | Optimal values of parameter<br>$\gamma$<br>in eq. (1) | Experimental value of the Fisher function $F$, eq. (3), $m=2$ | Critical value of Fisher function $F_c(l, n-2)$ for the confidence level 0.001, [29] | $F/F_c$ |
| --- | --- | --- | --- | --- | --- | --- | --- | --- |
| 2020, Japan | 316 | -0.4956 | 0.45 | 110.7141 | -1.3457 | 102.24 | 10.8 | 9.47 |
| 2021, Japan | 365 | -0.9903 | 0.33 | 944.4762 | -1.2316 | 18431.73 | 10.8 | 1706.6 |
| 2022, Japan | 100 | -0.9808 | 0.00 | 918.3058 | -1.2622 | 2483.54 | 11.6 | 214.1 |
| 2020, USA | 299 | -0.2603 | 4.14 | 574.36 | -0.6095 | 21.58 | 10.8 | 1.998 |
| 2021, USA | 365 | -0.9642 | 0.00 | 10222.8 | -1.3957 | 4796.96 | 10.8 | 444.2 |
| 2022, USA | 95 | -0.9962 | 3.27 | 5070.4 | -1.2371 | 12328.36 | 11.6 | 1062.8 |

**Fig. 5.**
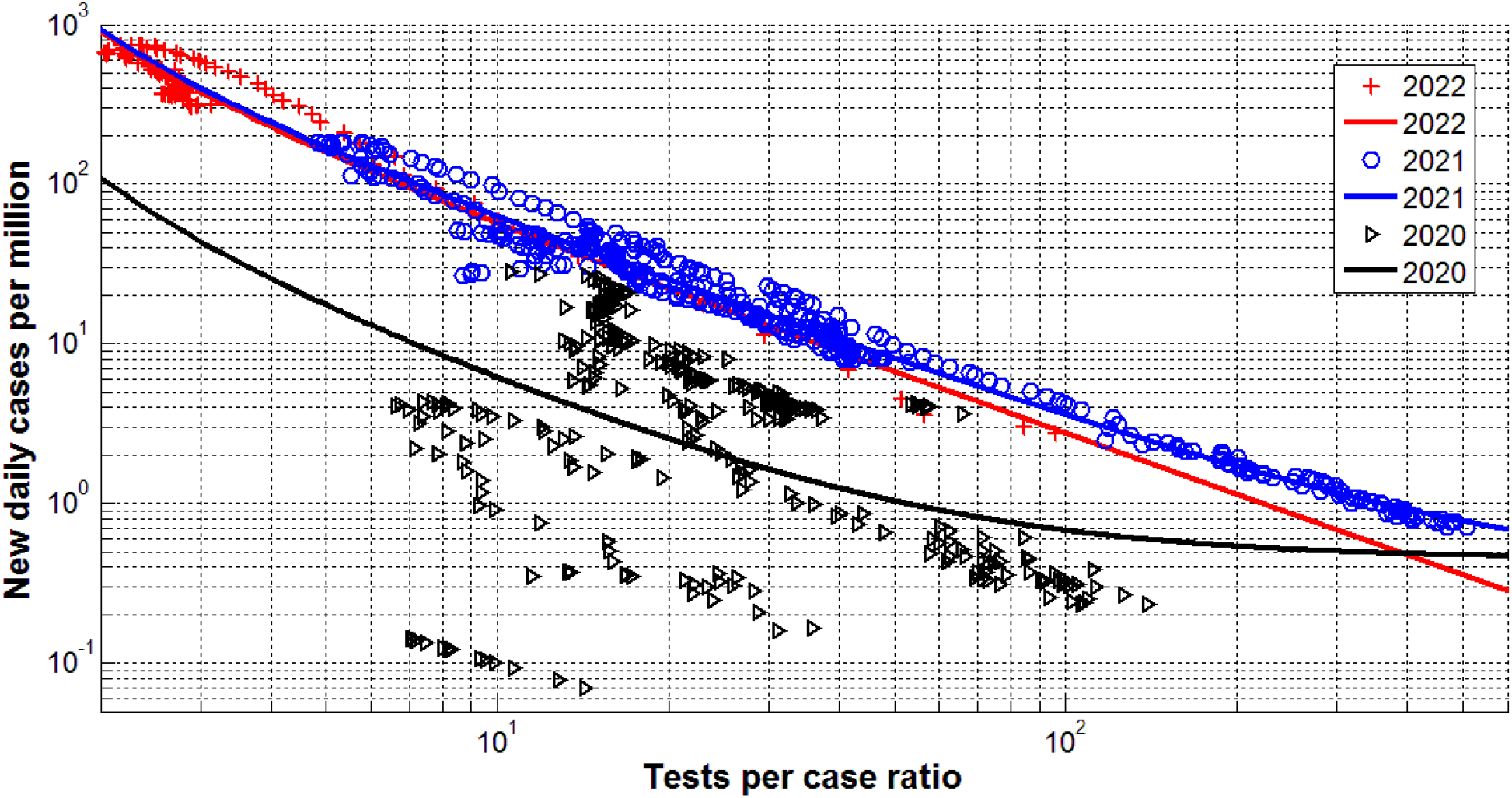
Averaged daily number of new cases per million (DCC) in Japan versus daily tests per case ratio (DTC=1000*TC/DCC) in 2020 (black), 2021 (blue) and 2022 (red). Markers represent the JHU data [27]. The results of non-linear regression (1) are shown by best fitting lines with the parameters listed in Table 1.

**Fig. 6.**
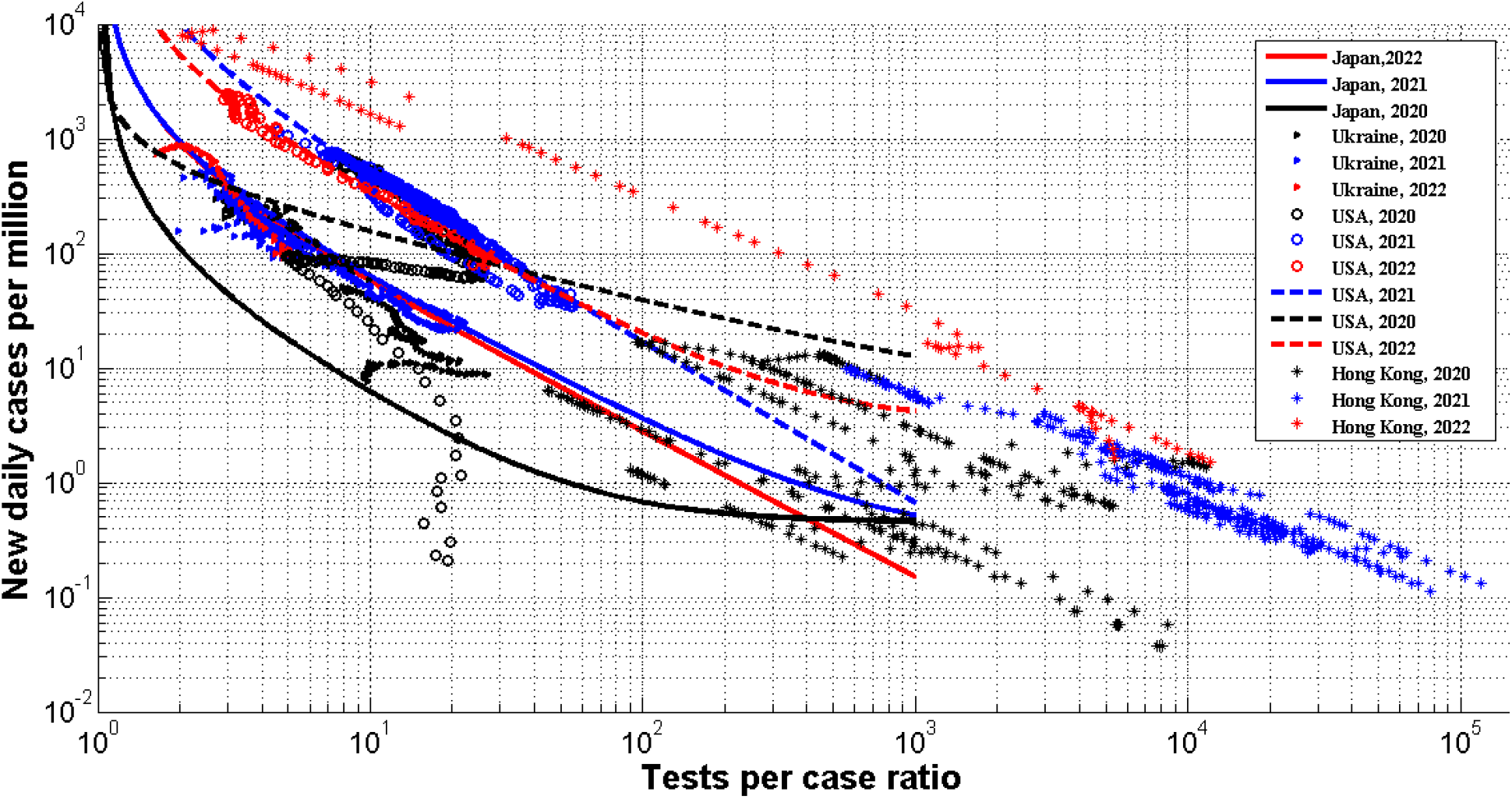
Averaged daily number of new cases per million (DCC) in Japan, Ukraine, USA, and Hong Kong versus daily tests per case ratio (DTC=1000*TC/DCC) in 2020 (black), 2021 (blue) and 2022 (red). ”Triangles” represent the JHU data [27] for Ukraine, “circles” – for USA, “stars” – Hong Kong. The results of non-linear regression (1) are shown by solid best fitting lines for Japan and dashed ones for USA.

Very interesting is the case of Hong Kong where the very high testing level was supported in 2020 and 2021 (see black and blue “stars” in Fig. 6). Due to this fact the numbers of new cases were less than 2 per million, i.e., the epidemic was controlled completely. The reduce of the tests per case ratio caused a huge epidemic wave with very high DCC values. In particular, the numbers of new cases per capita became even higher than at maximum level in the USA (compare red “stars” and “circles” in Fig. 6).

The relative characteristics in Hong Kong versus time in 2022 are shown in Fig. 7 (dashed lines). The daily number of new cases started to increase after January 18 (see the blue dashed line). Since the number of tests remained constant (see the black dashed line), the daily tests per case ratio diminished drastically (the red dashed line). After some increase of the number of tests (period of January 30 to February 5), the decrease of the test per case ratio stopped, and some stabilization of the number of cases is visible (the blue dashed line). But probably it was already not enough to control the epidemic outbreak, especially after fixing the number of tests. As a result, the drastic increase of the number of cases is visible for the period of February 6 to February 24 (see the blue dashed line). This increase has accelerated after the decrease of the number of tests in the end of February.

**Fig. 7.**
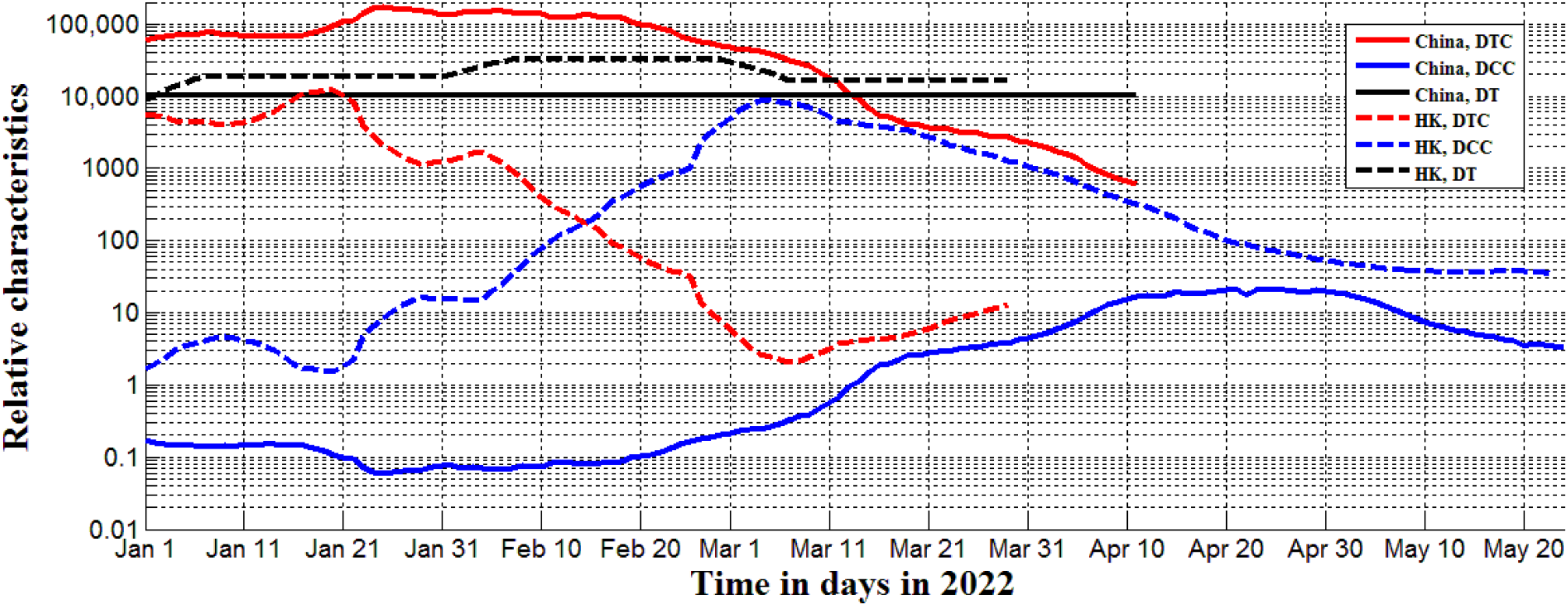
Pandemic dynamics and testing levels in mainland China (solid lines) and Hong Kong (HK, dashed lines) in 2022. Blue lines represents the smoothed daily numbers of new cases DCC, black ones – smoothed daily numbers of tests per million DT=1000*TC (the JHU datasets [27] published on May 26, 2022). Red lines shows the calculated daily tests per case ratio DTC=1000*TC/DCC.

The daily numbers of tests per capita in mainland China was constant in 2022 and lower than in Hong Kong (see solid and dashed black lines in Fig. 7). Due to much lower number of cases, the test per case ratio was much higher in mainland China (compare red solid and dashed lines in Fig. 7). Even after starting the new wave in February 2022, the DTC values in China did not drop under 500 (see the solid red line). This fact allowed controlling the pandemic. The daily number of new cases per million were lower than 20 in China (see the blue solid line).

## Discussion

To have more evidences that the increase of the daily tests per case ratio diminishes the DCC values, we have applied the non-linear regression (1) to the pandemic characteristics in Europe, Africa and some other countries listed in [24, 35]. The figures correspond to the same day – February 1, 2022 - and were extracted from the JHU tables [27] (version corresponding to February 6, 2022). The results of calculations are shown in Table 2 and Fig. 8. For Europe and complete dataset listed in [24] the non-linear correlation (1) is supported at the confidence level 0.025 (see first two rows in Table 2 and blue lines in Fig. 8). In the case of Africa, the confidence level even higher than 0.001.

**Table 2.**
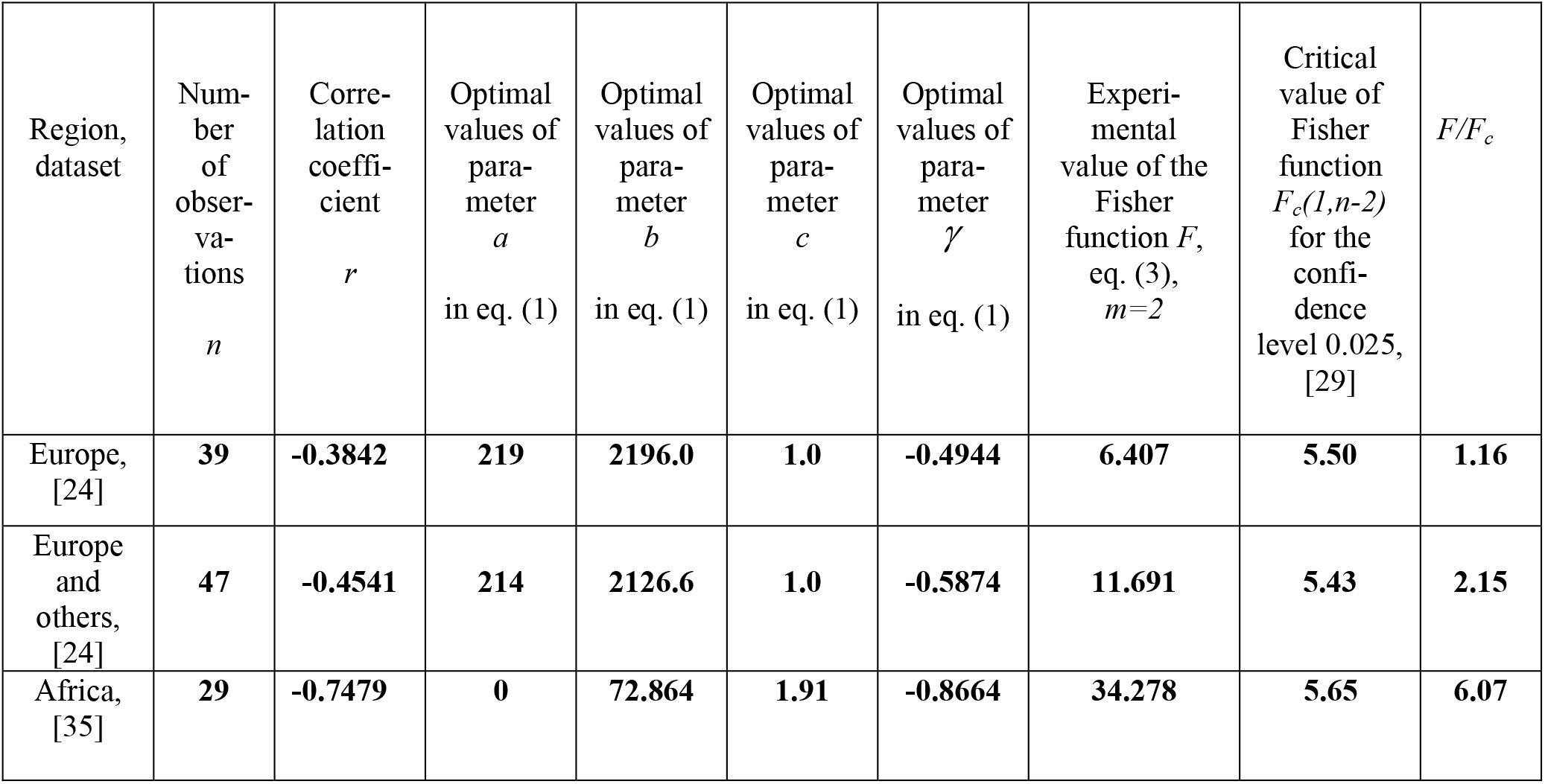
Results of non-linear regression DCC on DTC for datasets listed in [24, 35] corresponding to February 1, 2022. Optimal values of parameters in eq. (1), correlation coefficients and the results of Fisher test applications.

**Fig. 8.**
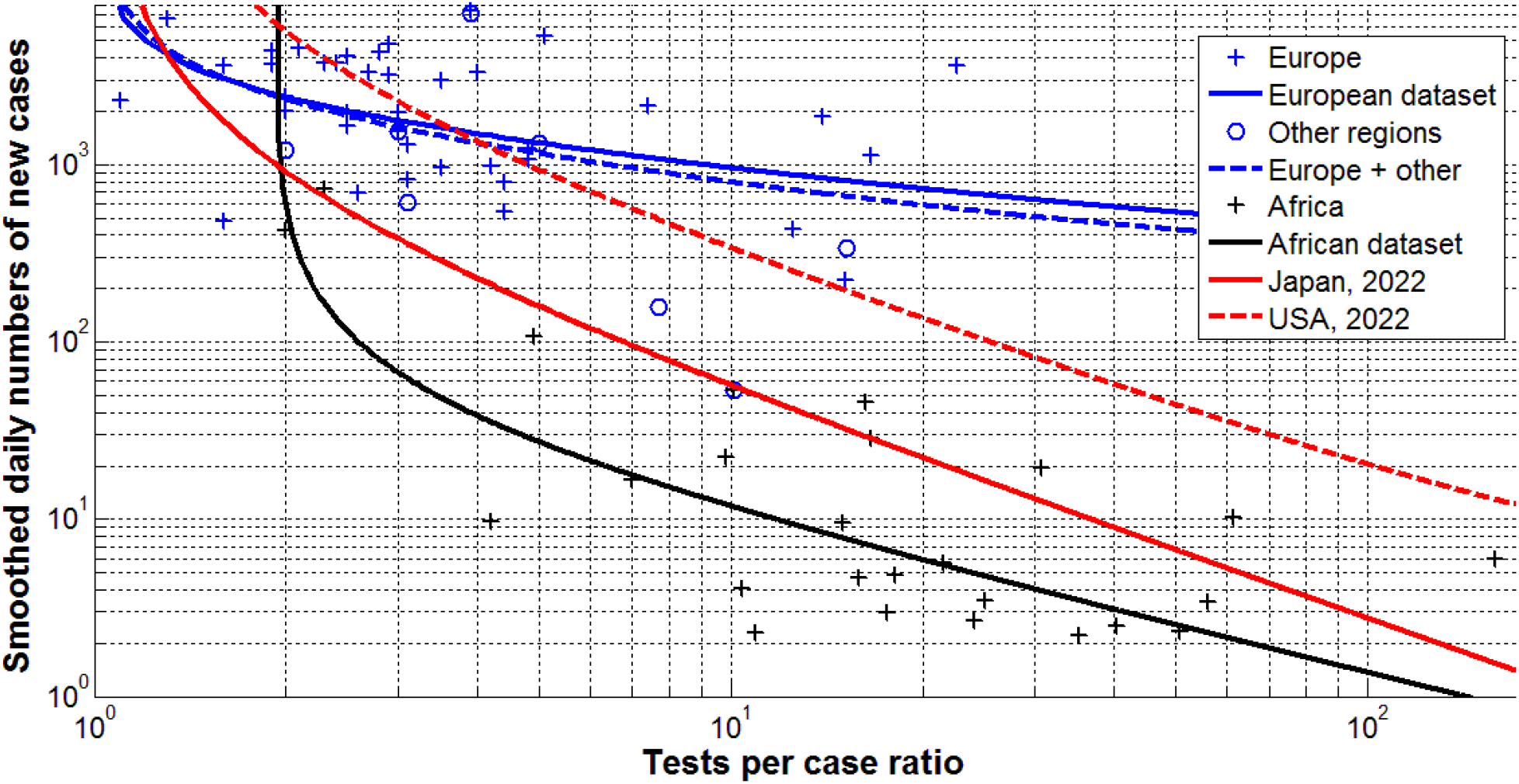
Averaged and smoothed daily number of new cases per million versus tests per case ratios (ATC and DTC, respectively) in European, African, and some other countries as of February 1, 2022. ”Crosses” represent the JHU data [27] for Europe (blue, [24]) and Africa (black, [35]); blue “circles” – other countries listed in [24]. The results of non-linear regression (according to eq. (1) and Table 2) are shown by best fitting lines (the solid blue one for Europe, the dashed blue - for Europe and some other countries listed in [24], the black - for African countries). Best fitting lines for DCC values in 2022 (according to Table 1) are shown in red (solid for Japan (eq. (4)) and dashed for USA).

The best fitting lines shown in Fig. 8 support the conclusion that the daily number of cases reduces with the increase of the daily tests per case ratio in Europe and some other countries listed in [24] (see blue lines). This trend is even more evident in Africa (see the black line), where the DTC vallues were in many cases higher than in European countries (compare black and blue “crosses” in Fig. 8). To compare with the daily numbers of new cases, the best fitting lines for Japan and USA in 2022 are shown by red solid and dashed lines, respectively. The corresponding optimal values of parameters were taken from Table 1. In particular, for Japan the best fitting non-linear equation can be written as follows:

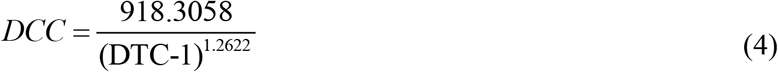

Fig. 8 demonstrates that the daily number of cases in Africa can be approximately 5 times lower than in Japan and 30 times lower than in USA at the same level of DTC. As of May 15, 2022 the accumulated numbers of COVID-19 cases per million in Africa (8,584), Japan (66,400) and USA (247,625), [27], follows these proportions. Probably the lower level of incomes and connected with this the lower mobility of population yielded very low relative level of infections in Africa. This hypothesis needs further investigations, but a very strong correlation between the per capita income and the accumulated numbers of cases per capita CC was revealed in [35].

Positive values of parameter *a* in eq. (1) show that new cases will appear even at very high testing levels, since DCC tends to *a* as DTC tends to infinity (at negative values of *γ*). For example, in 2022 the averaged daily number of new cases per million in USA cannot be less than 3.27 even at very high testing level (see the last row of Table 1). The estimation for Japan is more optimistic. Eq. (4) demonstrates that in 2022 the numbers of new cases in Japan could be reduced to zero at very high testing levels. For example, if the test per case ratio exceeds 500, the average daily number of new cases per million in Japan will be less than 0.36. This testing level was registered in 2021 with the corresponding number of daily deaths around 0.01 per million (see Fig. 4). The increase of the tests per case ratio in 2022 needs additional resources, since the number of new cases is still very high. Nevertheless, it could be an effective tool to control the pandemic in Japan and other countries. For this purpose the self testing systems can be accumulated during the periods between pandemic waves and than be used immediately after the beginning of new waves (in order to reduce the load on laboratories).

## Conclusions

The COVID-19 pandemic dynamics in 2020, 2021 and 2022 has been compared for Japan, Ukraine, USA, Hong Kong, mainland China, European and African countries. Some seasonal correlations for Japan were revealed. The presented results indicate that vaccinations do not prevent new infections, but vaccinated persons are less likely to die. Non-linear correlation analysis has revealed the close links between the smoothed daily numbers of new cases and smoothed daily values of the test per case ratio. To decrease the number of new cases and control the pandemic effectively, the tests per case ratio has to be significantly increased.

## Data Availability

All data produced in the present work are contained in the manuscript

## Acknowledgement

The author is very grateful to Professor Hiroshi Nishiura (Kyoto University School of Public Health) for his support and useful discussions. The author thanks Oleksii Rodionov for his help in collecting and processing data.

